# Elucidating distinct and common fMRI-complexity patterns in pre-adolescent children with Attention-Deficit/Hyperactivity Disorder, Oppositional Defiant Disorder, and Obsessive-Compulsive Disorder diagnoses

**DOI:** 10.1101/2025.01.17.25320748

**Authors:** Ru Zhang, Steven Cen, Dilmini Wijesinghe, Leon Aksman, Stuart B. Murray, Christina J. Duval, Danny J.J. Wang, Kay Jann

## Abstract

**Importance:** The pathophysiology of ADHD is complicated by high rates of psychiatric comorbidities, thus delineating unique versus shared functional brain perturbations is critical in elucidating illness pathophysiology.

**Objective:** To investigate resting-state fMRI (rsfMRI)-complexity alterations among children with ADHD, oppositional defiant disorder (ODD), and obsessive-compulsive disorder (OCD), respectively, and comorbid ADHD, ODD, and OCD, within the cool and hot executive function (EF) networks.

**Design:** We leveraged baseline data (wave 0) from the Adolescent Brain and Cognitive Development (ABCD) Study.

**Setting:** The data was collected between September 2016 and September 2019 from 21 sites in the USA.

**Participants:** Children who singularly met all DSM-5 behavioral criteria for ADHD (*N* = 61), ODD (*N* = 38), and OCD (*N* = 48), respectively, were extracted, alongside children with comorbid ADHD, ODD, OCD, and/or other psychiatric diagnoses (*N* = 833). A control sample of age-, sex-, and developmentally-matched children was also extracted (*N* = 269).

**Main Outcomes and Measures:** Voxel-wise sample entropy (SampEn) was computed using the LOFT Complexity Toolbox. Mean SampEn within all regions of the EF networks was calculated for each participant and hierarchical models with Generalized Estimating Equations compared SampEn of comorbid-free and comorbid ADHD, ODD, and OCD within the EF networks.

**Results:** SampEn was reduced in comorbid-free ADHD and ODD in overlapping regions of both EF networks, including the bilateral superior frontal gyrus, anterior/posterior cingulate gyrus, and bilateral caudate (Wald statistic = 5.682 to 10.798, *p* < 0.05 & BH corrected), with ADHD additionally affected in the right inferior/middle frontal gyrus and bilateral frontal orbital cortex (Wald statistic = 7.231 to 9.420, *p* < 0.05 & BH corrected). Among comorbid presentations, the additional presence of ADHD symptomatology was associated with significantly lower SampEn in every region of interest (*z* = -3.973 to -2.235, *p* < 0.05 & BH corrected).

**Conclusions and Relevance:** ADHD and ODD shared common impairments underlying the EF networks in the comorbid-free presentations, with ADHD showing more widespread complexity reduction. When ADHD co-occurred with other psychiatric disorders, the reduction in SampEn extended beyond the regions affected in comorbid-free ADHD, indicating that comorbidities amplify neural complexity deficits.

## Introduction

Attention-deficit/hyperactivity disorder (ADHD) is a childhood neurodevelopmental disorder that affects 11.3% of children and adolescents and approximately 4.4% of adults in the United States.^1–3^ Although numerous studies have been conducted on ADHD, the pathophysiology of the disorder is still not fully understood, especially due to the high rate of comorbidities. Two commonly observed comorbid presentations are obsessive-compulsive disorder (OCD) and oppositional defiant disorder (ODD).

All three disorders are characterized by deficits in executive function (EF). EF can be distinguished into hot and cool EF,^4^ whereby cool EF refers to goal-directed and problem-solving behaviors, as well as self-regulation, not involving motivational or affective aspects, and hot EF is characterized by motivational and affective aspects of cognitive processing. ADHD is characterized predominantly by structural and functional brain alterations in the cool EF system, whereas ODD is associated with abnormalities in the hot EF system, and OCD may include alterations in both.^5–7^

Nonlinear analyses from functional MRI that characterize neural signal complexity have been proposed as measures for information processing capacity of brain areas and networks,^8–11^ or indices of pathological brain function.^12–14^ Sample entropy (SampEn) has attracted considerable attention in complexity analysis.^15^ It captures signal regularity that is sensitive to subtle, dynamic changes in the brain, often overlooked by the conventional connectivity or network-based approaches.^16–22^ Few studies have investigated the complexity of resting-state fMRI (rsfMRI) data in ADHD, two for adult ADHD^23, 24^ and one for pre-adolescent ADHD,^25^ and all reported reduced complexity in frontal cortex. Complexity studies in OCD have been focused on adult participants and the results are diverse.^26–28^ No study has investigated complexity of ODD.

The current study represents the first attempt to explore functional brain alterations in ADHD, ODD, and OCD, both in the comorbid-free and comorbid presentations. We examined SampEn of ADHD, ODD, and OCD in the cool and hot EF networks when they were comorbid-free disorders, as well as when these symptomatologies co-occurred. We hypothesized that: (1) SampEn for comorbid-free ADHD, ODD, and OCD would be lower when compared to healthy controls, and (2) SampEn in the presence of ADHD, ODD, and OCD symptomatologies would be lower than those in absence.

## Methods

### Participants

We leveraged the baseline demographic, clinical, T1 structural, and rsfMRI data from release 3.0 of Adolescent Brain and Cognitive Development (ABCD) Study.^29^ The ABCD baseline included 11878 participants in total, and each was evaluated based on 144 psychiatric diagnoses (see Figure S1). 1087 of them were diagnosed with ADHD. More details about the diagnosis of ADHD in ABCD release 3.0 are in the supplementary material.

We divided the whole ADHD sample into 8 groups, i.e., (i) ADHD, (ii) ADHD with comorbid ODD, (iii) ADHD with comorbid OCD, (iv) ADHD with comorbid ODD and OCD, (v) ADHD with another psychiatric comorbidity, (vi) ADHD with comorbid ODD and another psychiatric comorbidity, (vii) ADHD with comorbid OCD and another psychiatric comorbidity, and (viii) ADHD with comorbid ODD, OCD, and another psychiatric comorbidity.

In addition, we extracted groups of comorbid-free ODD and comorbid-free OCD. We also pseudorandomly selected a healthy control group, with matched distributions in youth pubertal development status (YPDS), sex, and age with the whole ADHD sample (*p* > 0.05). We additionally required each participant to have T1 structural data and at least two complete rsfMRI scans. Participants with average framewise displacement (FD) > 0.2 mm in any rsfMRI scan run were excluded. The comorbid-free ADHD, ODD, and OCD matched with the healthy controls in terms of sex, age, YPDS, and average FD (avgFD) over two rsfMRI scan runs, and scan-site (Table 1). The other ADHD groups matched with the healthy controls in terms of sex, age, and YPDS but not avgFD and scan-site (*p* < 0.05, see Table 2).

**Table 1.**
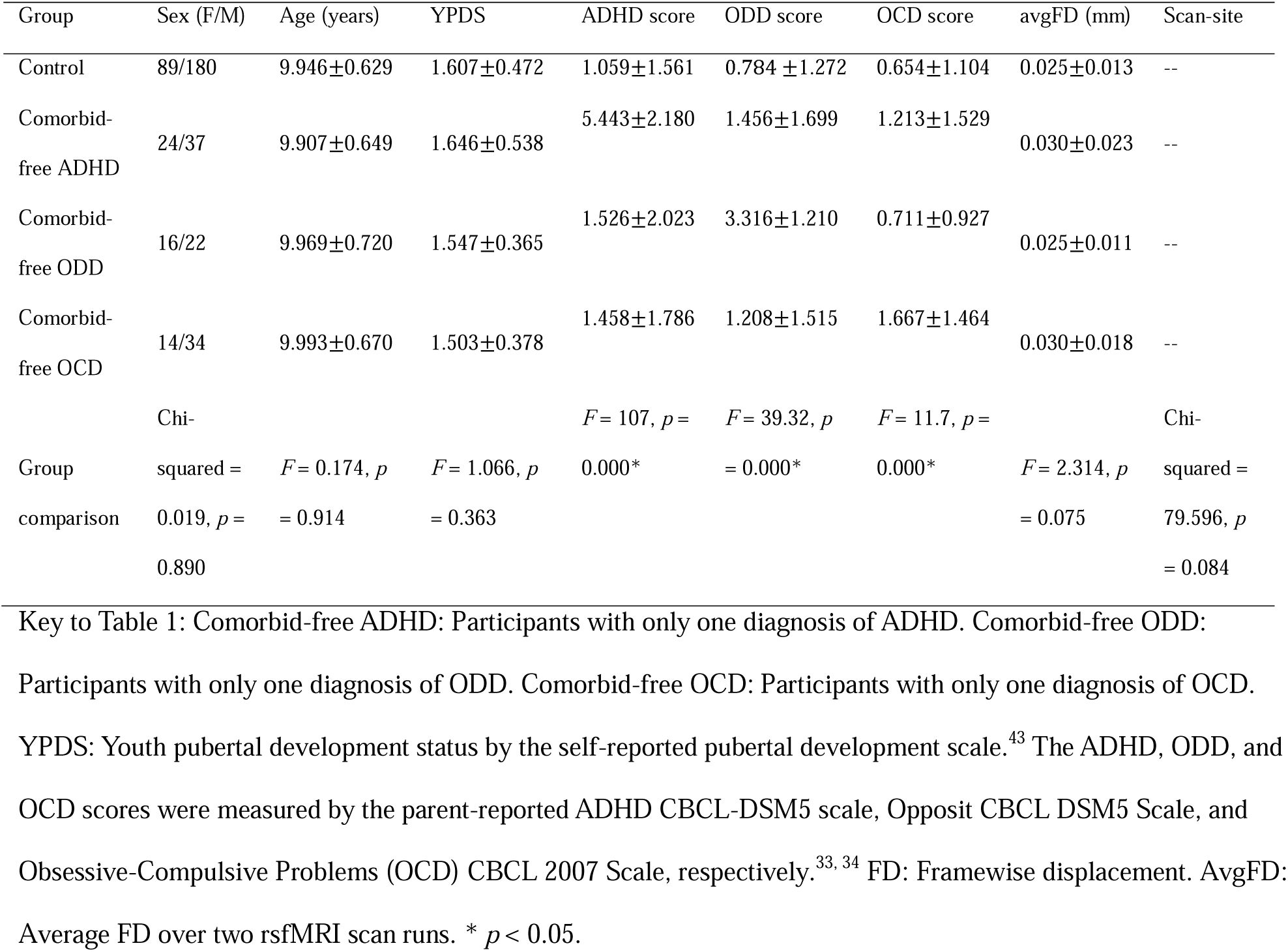
An overview of sample characteristics delineated by group, for the analysis of comorbid-free ADHD, ODD, and OCD.

**Table 2.**
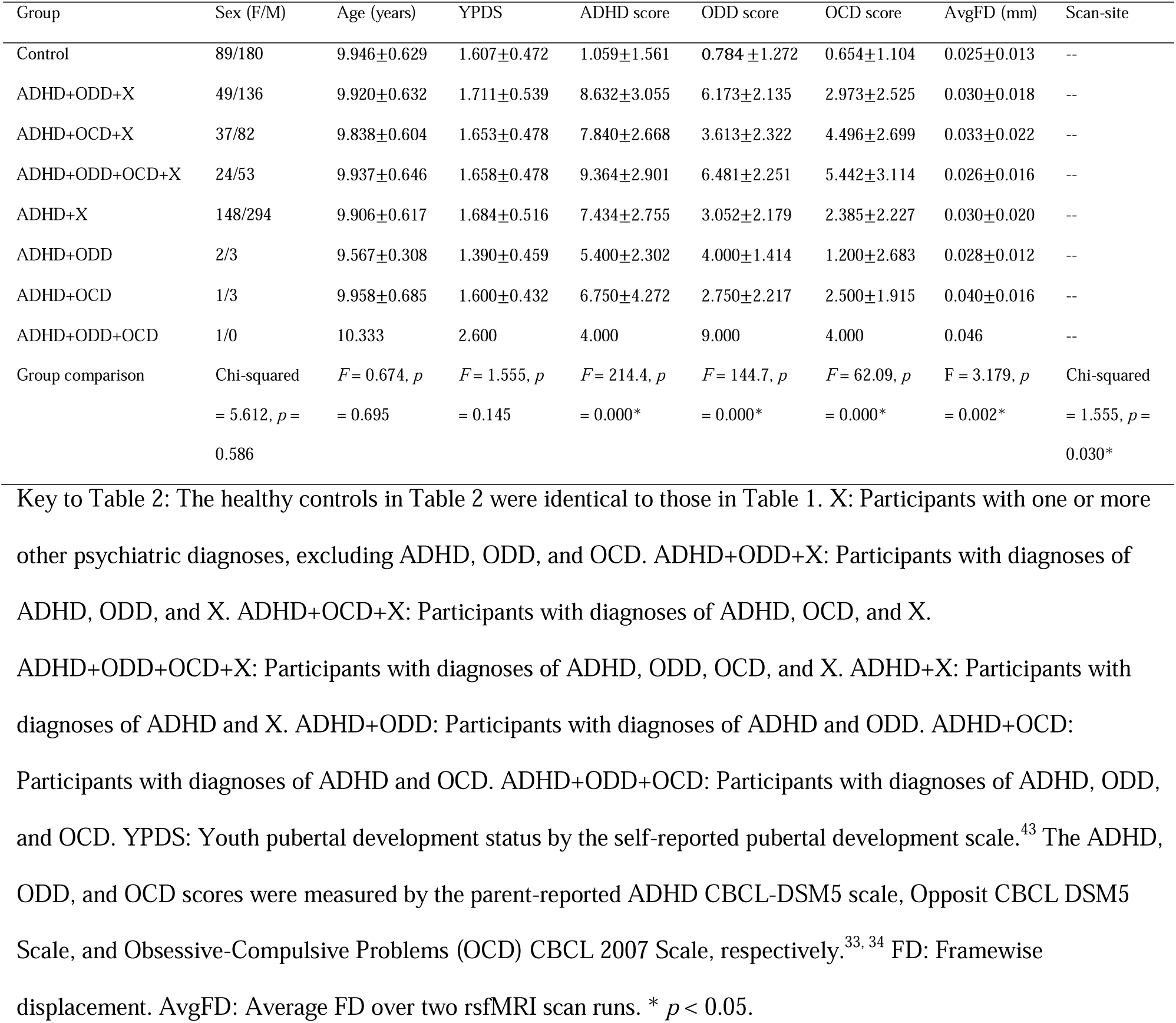
An overview of sample characteristics delineated by group, for the analysis of comorbid ADHD, ODD, and OCD.

### Data Analyses

#### Complexity Analysis

Data preprocessing was performed in CONN toolbox (details in the supplementary material). Voxel-wise SampEn was computed using the LOFT Complexity Toolbox. SampEn is defined as the natural logarithm of the conditional probability that a pattern length of *m* points will repeat itself, excluding self-matches, for *m* + 1 points within a tolerance of *r* in a time series of length *N.*^15^ We set the pattern length *m* = 2 and the sensitivity threshold *r* = 0.3 in the current study.^30^ SampEn was computed for each voxel for each rsfMRI time series and then averaged over the two rsfMRI runs for each participant.

Individual mean SampEn within each region-of-interest (ROI) of the two networks was calculated: ROIs based on the Harvard-Oxford atlas^31^ for the cool EF network included the superior frontal gyrus, middle frontal gyrus, inferior frontal gyrus, anterior cingulate cortex, and hippocampus.^4^ Hot EF network included the amygdala, accumbens, caudate, putamen, anterior cingulate cortex, frontal medial cortex, frontal orbital cortex, and posterior cingulate gyrus.^4^ Mean SampEn of each network was also calculated.

#### Analysis for Comorbid-free ADHD, ODD, and OCD

Hierarchical models with Generalized Estimating Equations (GEE) were implemented to investigate SampEn of the comorbid-free ADHD, ODD, and OCD vs. healthy controls using avgFD as a covariate and scan-site as a clustering variable for (1) each EF network as a whole and (2) each ROI in each EF network separately (see information of the participants in Table 1).

All the models tested in the current article included network-wise and ROI analyses. Benjamini-Hochberg (BH) correction^32^ with false discovery rate *q* < 0.05 was performed for all the analyses in this article.

#### Analysis for the Whole ADHD Sample

Hierarchical models with GEE were implemented to investigate SampEn of the whole ADHD sample vs. healthy controls with avgFD as a covariate and scan-site as the clustering factor. Here, the whole ADHD sample included individuals with ADHD with all kinds of comorbidities (Table 2) and comorbid-free ADHD (Table 1).

#### Analysis for Comorbid ADHD, ODD, and OCD

Hierarchical models with GEE were used to investigate SampEn affected by ADHD symptomatology (presence vs. absence), ODD symptomatology (presence vs. absence), and OCD symptomatology (presence vs. absence). Here ADHD, ODD, and OCD symptomatologies were treated as the independent variables, other symptomatologies (presence vs. absence) and avgFD as covariates, and scan-site as the clustering factor. Only participants displayed in Table 2 were used in this analysis.

#### Secondary Analyses

##### Effects of Number of Comorbidities

Hierarchical models with GEE were used to evaluate the association between SampEn and the number of comorbidities with avgFD as a covariate and scan-site as the clustering factor. Note the whole ADHD sample was included in this analysis but all the healthy controls were excluded.

##### Association between Complexity and Symptom Severity

The ADHD, ODD, and OCD scores were measured by the parent-reported ADHD CBCL-DSM5 scale, Opposit CBCL DSM5 Scale, and Obsessive-Compulsive Problems (OCD) CBCL 2007 Scale, respectively^33, 34^. Hierarchical models with GEE were used to evaluate the association between SampEn and the ADHD score within the whole ADHD sample using avgFD as a covariate and scan-site as the clustering factor. The same analysis was replicated for the ODD score within the whole ODD sample and the OCD score within the whole OCD sample.

##### Medication Effects

Hierarchical models with GEE examined the main effect of medication (medicated vs. unmedicated) within the whole ADHD sample using avgFD as a covariate and scan-site as the clustering factor. Please refer to Table S1 in the supplementary material for the list of prescriptions. The ADHD participants who consumed at least one prescription were categorized as medicated.

## Results

### Clinical Results

There was no statistical difference in terms of sex (χ^2^= 0.019, *p* > 0.05), age (*F* = 0.174, *p* > 0.05), YPDS (*F* = 1.066, *p* > 0.05), avgFD (*F* = 2.314, *p* > 0.05), and scan-site (χ^2^= 79.596, *p* > 0.05) when comparing the comorbid-free ADHD, ODD, OCD, and the healthy controls (Table 1). There was no statistical difference in terms of sex (χ^2^= 5.612, *p* > 0.05), age (*F* = 0.674, *p* > 0.05), and YPDS (*F* =1.555, *p* > 0.05), but not avgFD (*F* = 3.179, *p* < 0.05) and scan-site (χ^2^= 1.555, *p* < 0.05) when comparing the comorbid ADHD groups and the controls (Table 2). More clinical results about symptom severity can be accessed in the supplementary material.

### Complexity Analysis

Mean SampEn for each subject in each group for the cool EF network and hot EF network is displayed in Figure 1.

**Figure 1.**
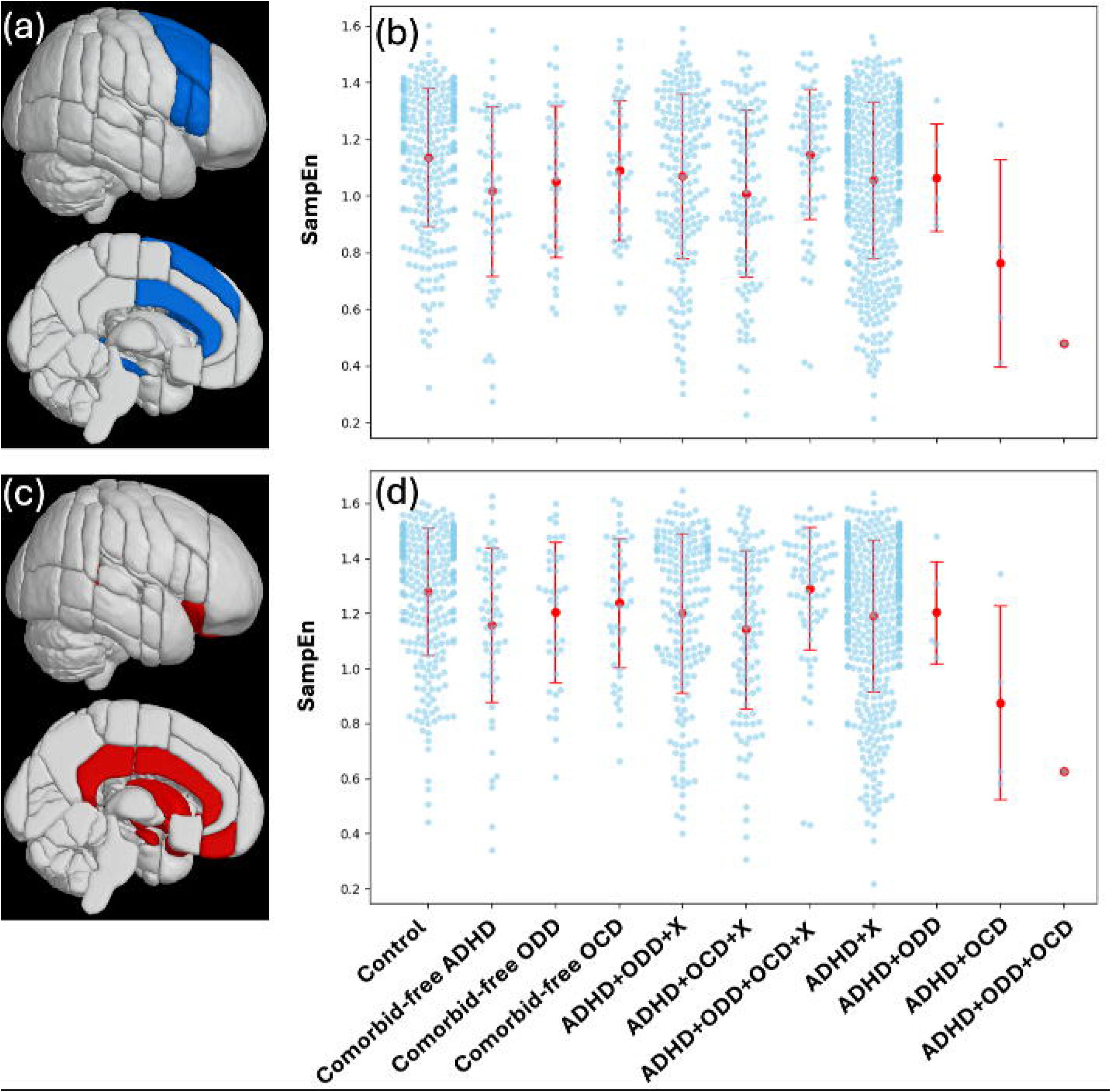
(a) The cool EF network. (b) Mean SampEn of the cool EF network for each subject, plotted against each group. The red dots and line segments display the means and standard deviations for the groups. (c) The hot EF network. (d) Mean SampEn of the hot EF network for each subject, plotted against each group. The red dots and line segments display the means and standard deviations for the groups.

### Analysis for Comorbid-free ADHD, ODD, and OCD

On the network level analysis, we found significant main effects of Group in each network (*p* < 0.05 & BH corrected). Post hoc tests indicated that comorbid-free ADHD and comorbid-free ODD had significantly lower SampEn than the controls in the cool EF network (Wald statistic = 6.920 & 5.371, *p* < 0.05 & BH corrected) and in the hot EF network (Wald statistic = 8.640 & 5.577, *p* < 0.05 & BH corrected). SampEn of comorbid-free OCD was not significantly different from the controls in either EF network.

For ROI analysis, we found main effects of Group in the bilateral superior frontal gyrus, right middle frontal gyrus, right inferior frontal gyrus, and anterior cingulate gyrus (χ^2^= 10.177 to 13.261, *p* < 0.05 & BH corrected) in the cool EF network, and in the anterior cingulate gyrus, posterior cingulate gyrus, bilateral frontal orbital cortex, and bilateral caudate in the hot EF network (χ^2^= 10.138 to 15.476, *p* < 0.05 & BH corrected). Post hoc tests indicated that (1) the comorbid-free ADHD had lower SampEn in the bilateral superior frontal gyrus, right middle frontal gyrus, right inferior frontal gyrus, and anterior cingulate gyrus in the cool EF network (Wald statistic = 6.355 to 9.420, *p* < 0.05 & BH corrected) and in the anterior cingulate gyrus, posterior cingulate gyrus, bilateral frontal orbital cortex, and bilateral caudate in the hot EF network relative to the controls (Wald statistic = 5.913 to 10.798, *p* < 0.05 & BH corrected); (2) the comorbid-free ODD had lower SampEn in the bilateral superior frontal gyrus and anterior cingulate gyrus in the cool EF network (Wald statistic = 6.650 to 7.518, *p* < 0.05 & BH corrected) and in the anterior cingulate gyrus, posterior cingulate gyrus, and bilateral caudate in the hot EF network compared to the controls (Wald statistic = 5.682 to 6.873, *p* < 0.05 & BH corrected); (3) the comorbid-free OCD and controls did not have significantly different SampEn in any ROI of any EF network (see Table 3 & Figure 2).

**Figure 2.**
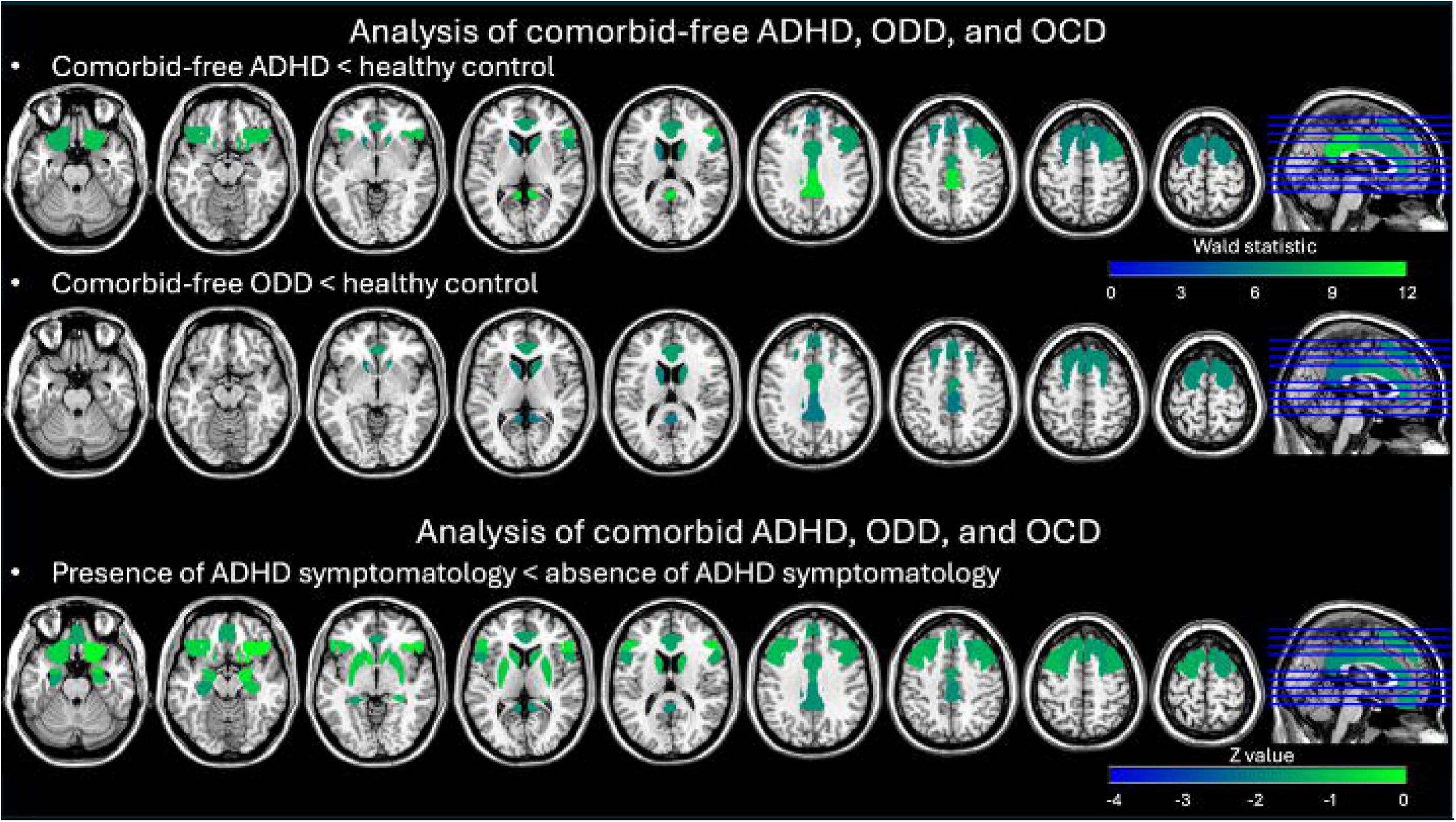
The significant results of analysis using hierarchical models with GEE for SampEn of ADHD, ODD, and OCD in comorbid-free and comorbid presentations. All the results present with *p* < 0.05 and BH correction. For the analysis of comorbid-free ADHD, ODD, and OCD, the model was SampEn = Group (comorbid-free ADHD, comorbid-free ODD, comorbid-free OCD vs. healthy controls) + avgFD, in which scan-site was a clustering factor. For the analysis of comorbid ADHD, ODD, and OCD, the model was SampEn = ADHD symptomatology (presence vs. absence) + ODD symptomatology (presence vs. absence) + OCD symptomatology (presence vs. absence) + X symptomatology (presence vs. absence) + avgFD, in which scan-site was a clustering factor.

**Table 3.**
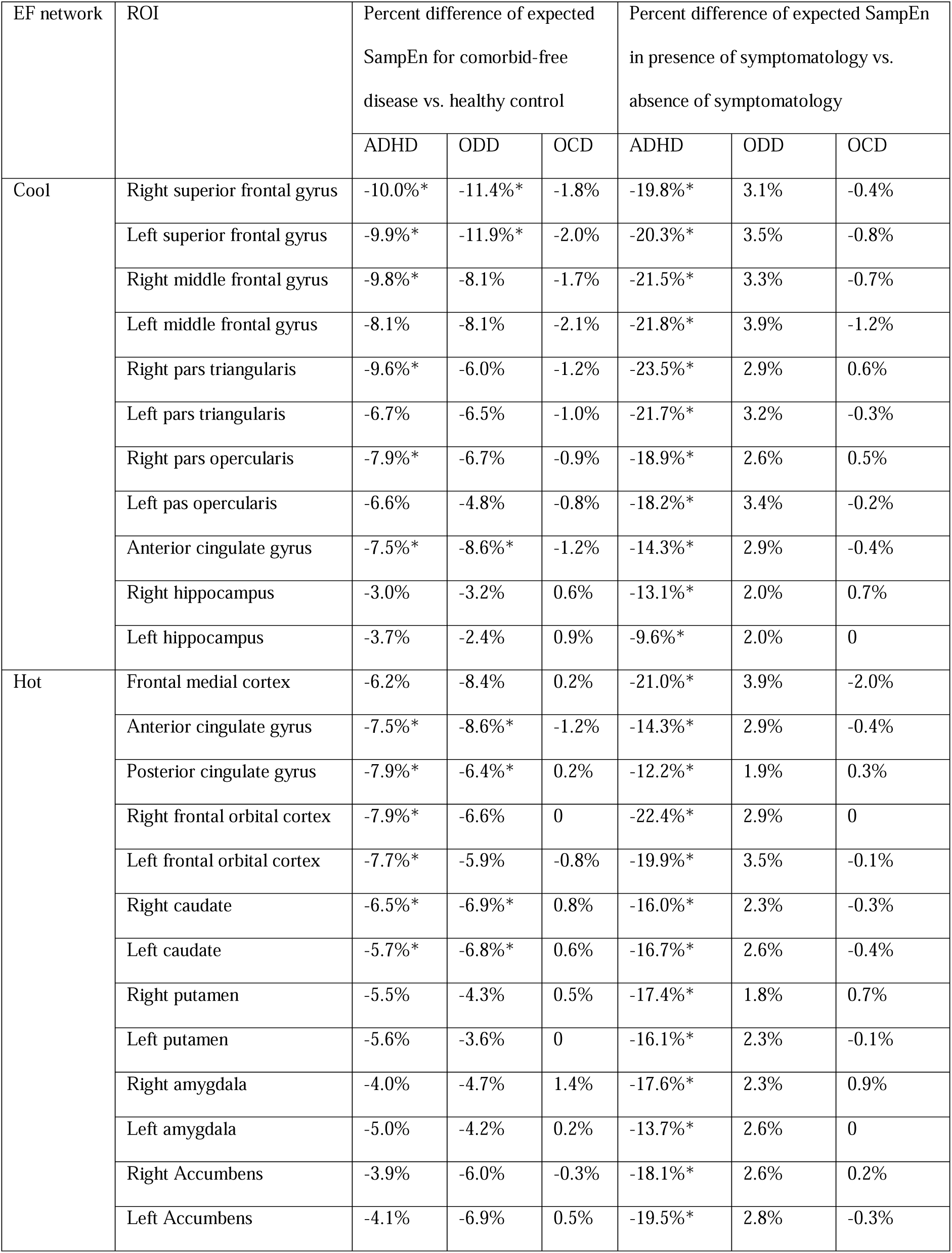
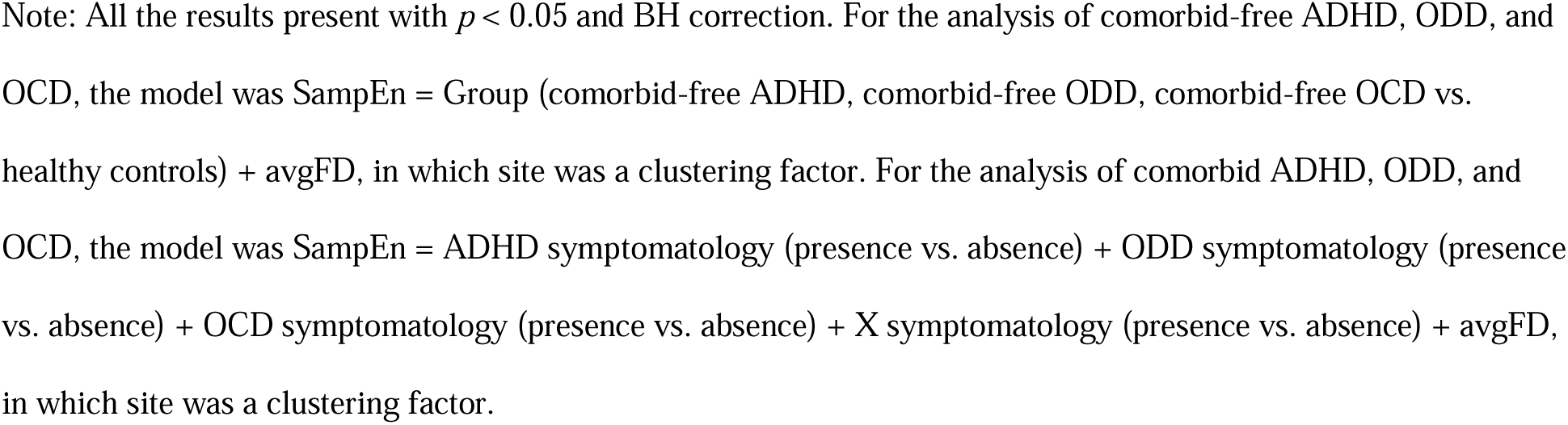
The significant results of analysis using hierarchical models with GEE for SampEn of ADHD, ODD, and OCD in comorbid-free and comorbid presentations.

| EF network | ROI | Percent difference of expected<br>SampEn for comorbid-free<br>disease vs. healthy control |  |  | Percent difference of expected SampEn<br>in presence of symptomatology vs.<br>absence of symptomatology |  |  |
| --- | --- | --- | --- | --- | --- | --- | --- |
|  |  | ADHD | ODD | OCD | ADHD | ODD | OCD |
| Cool | Right superior frontal gyrus | -10.0% * | -11.4% * | -1.8% | -19.8% * | 3.1% | -0.4% |
|  | Left superior frontal gyrus | -9.9% * | -11.9% * | -2.0% | -20.3% * | 3.5% | -0.8% |
|  | Right middle frontal gyrus | -9.8% * | -8.1% | -1.7% | -21.5% * | 3.3% | -0.7% |
|  | Left middle frontal gyrus | -8.1% | -8.1% | -2.1% | -21.8% * | 3.9% | -1.2% |
|  | Right pars triangularis | -9.6% * | -6.0% | -1.2% | -23.5% * | 2.9% | 0.6% |
|  | Left pars triangularis | -6.7% | -6.5% | -1.0% | -21.7% * | 3.2% | -0.3% |
|  | Right pars opercularis | -7.9% * | -6.7% | -0.9% | -18.9% * | 2.6% | 0.5% |
|  | Left pas opercularis | -6.6% | -4.8% | -0.8% | -18.2% * | 3.4% | -0.2% |
|  | Anterior cingulate gyrus | -7.5% * | -8.6% * | -1.2% | -14.3% * | 2.9% | -0.4% |
|  | Right hippocampus | -3.0% | -3.2% | 0.6% | -13.1% * | 2.0% | 0.7% |
|  | Left hippocampus | -3.7% | -2.4% | 0.9% | -9.6% * | 2.0% | 0 |
| Hot | Frontal medial cortex | -6.2% | -8.4% | 0.2% | -21.0% * | 3.9% | -2.0% |
|  | Anterior cingulate gyrus | -7.5% * | -8.6% * | -1.2% | -14.3% * | 2.9% | -0.4% |
|  | Posterior cingulate gyrus | -7.9% * | -6.4% * | 0.2% | -12.2% * | 1.9% | 0.3% |
|  | Right frontal orbital cortex | -7.9% * | -6.6% | 0 | -22.4% * | 2.9% | 0 |
|  | Left frontal orbital cortex | -7.7% * | -5.9% | -0.8% | -19.9% * | 3.5% | -0.1% |
|  | Right caudate | -6.5% * | -6.9% * | 0.8% | -16.0% * | 2.3% | -0.3% |
|  | Left caudate | -5.7% * | -6.8% * | 0.6% | -16.7% * | 2.6% | -0.4% |
|  | Right putamen | -5.5% | -4.3% | 0.5% | -17.4% * | 1.8% | 0.7% |
|  | Left putamen | -5.6% | -3.6% | 0 | -16.1% * | 2.3% | -0.1% |
|  | Right amygdala | -4.0% | -4.7% | 1.4% | -17.6% * | 2.3% | 0.9% |
|  | Left amygdala | -5.0% | -4.2% | 0.2% | -13.7% * | 2.6% | 0 |
|  | Right Accumbens | -3.9% | -6.0% | -0.3% | -18.1% * | 2.6% | 0.2% |
|  | Left Accumbens | -4.1% | -6.9% | 0.5% | -19.5% * | 2.8% | -0.3% |

Note: All the results present with $p < 0.05$ and BH correction. For the analysis of comorbid-free ADHD, ODD, and OCD, the model was $\text{SampEn} = \text{Group (comorbid-free ADHD, comorbid-free ODD, comorbid-free OCD vs. healthy controls)} + \text{avgFD}$ , in which site was a clustering factor. For the analysis of comorbid ADHD, ODD, and OCD, the model was $\text{SampEn} = \text{ADHD symptomatology (presence vs. absence)} + \text{ODD symptomatology (presence vs. absence)} + \text{OCD symptomatology (presence vs. absence)} + \text{X symptomatology (presence vs. absence)} + \text{avgFD}$ , in which site was a clustering factor.

### Analysis for the Whole ADHD Sample

The whole ADHD sample showed significantly lower SampEn than the healthy controls in both EF networks (z = -2.881 & -3.268, *p* < 0.05 & BH corrected). In addition, the whole ADHD sample had significantly lower SampEn than the healthy controls in every ROI in each EF network (*z* = -3.617 to -2.550, *p* < 0.05 & BH corrected).

### Analysis for Comorbid ADHD, ODD, and OCD

On the network level, the presence of ADHD symptomatology was associated with significantly lower SampEn in both EF networks (z = -2.824 & -3.139, *p* < 0.05 & BH corrected). There was no significant difference between the presence vs. absence of ODD symptomatology and between the presence vs. absence of OCD symptomatology in each EF network.

For ROI analysis, we found that the presence of ADHD symptomatology was associated with significantly lower SampEn in every ROI in each EF network (*z* = -3.973 to -2.235, *p* < 0.05 & BH corrected). There was no significant difference between the presence vs. absence of ODD symptomatology and between the presence vs. absence of OCD symptomatology in any ROI.

### Secondary Analyses

#### Effects of Number of Comorbidities

Mean SampEn of each EF network was plotted against the number of comorbidities for each subject in Figure S1. We found no significant association between SampEn and the number of comorbidities in any EF network and in any ROI of any EF network.

#### Association between Complexity and Symptom Severity

No significant associations were found between SampEn and ADHD scores in any EF network or ROI for the whole ADHD sample. Similarly, no significant associations were observed between SampEn and ODD scores in the whole ODD sample, or between SampEn and OCD scores in the whole OCD sample. Notably however, symptom severity was higher in comorbid cases than in comorbid-free cases (Supplemental Material).

#### Medication Effects

There was no significant difference in SampEn between the medicated and non-medicated ADHD participants in any EF network or in any ROI.

## Discussion

We demonstrated that individuals with comorbid-free ADHD exhibited significantly reduced SampEn across many of the ROIs in both cold and hot EF networks, while comorbid-free ODD showed reduced SampEn in a subset of overlapping ROIs. When ADHD co-occurred with various comorbidities, we showed the reduction in SampEn impacted all ROIs within both EF networks, suggesting a cumulative disease burden and more severe deficits. Our analysis further revealed that reduced SampEn observed in the whole ADHD sample is primarily driven by ADHD symptomatology rather than the severity of comorbidities.

Previous studies have reported reduced rsfMRI complexity in ADHD,^23–25^ which were corroborated by our findings using a larger sample and accounting for comorbidities. Specifically, we observed reduced SampEn in ADHD with comorbid pathologies as well as in comorbid-free ADHD within both cool and hot EF networks.

Existing literature on task-based fMRI studies showed that ADHD is characterized predominantly by functional alterations in the cool EF system,^6^ while previous rsfMRI-based meta-analyses reported ADHD featured altered functional connectivity between regions in both cool and hot EF networks.^35–37^ Notably, in our previous study,^25^ we reported decreased complexity in association with altered functional connectivity in the same cohort of comorbid-free ADHD children but compared to a completely distinct set of matched control subjects, thus providing evidence of reliability of this finding and a relation between complexity and connectivity in brain networks.

In comorbid-free ODD, our study revealed reduced SampEn within both EF networks, although fewer regions were affected compared to comorbid-free ADHD. ODD has primarily been associated with abnormalities in the hot EF system and most functional imaging studies focused on this EF system,^5^ there is sparse evidence for cold EF deficits.^38^ Our study is the first to investigate the complexity of brain function in ODD and provides supportive evidence for impairments not only in the hot EF network but also the cold EF network. While the cold EF network is responsible for cognitive control, deficits in this area may also exacerbate dysfunctions in hot EF, which involve emotional and reward-based decision-making. The current observation highlights the need to include both networks to understand disease-specific and potential transdiagnostic deficits in ODD.

Research on OCD complexity has so far exclusively focused on adult populations, with studies reporting both increased and decreased complexity.^26–28^ Similarly, structural and functional neuroimaging findings in adult populations with OCD have revealed varied findings that lack generalizability.^39^ Neuroimaging in pediatric OCD is a growing field, and findings across studies and modalities vary with most commonly observed abnormalities in cortico-striatal loops.^40^ While our study is the first to investigate the complexity of pediatric OCD, we did not find significantly different SampEn in comorbid-free OCD compared to the controls in the EF networks. This however does not preclude differences on an individual level or other brain networks. Additionally, only a portion of youth with OCD persist into OCD in adulthood, which could indicate different pathophysiological subgroups, thus warranting further research to elucidate the neurophysiology and developmental trajectories of OCD.

Our analysis of the entire ADHD sample suggests more widespread deficits, including all ROIs within both EF networks, when comorbidities are present. However, there was no significant association between SampEn and the number of comorbidities. When the whole ADHD sample was divided into subgroups with varying comorbidities (ODD, OCD, and other psychiatric disorders), ADHD symptomatology remained the dominant factor in reducing SampEn. In contrast, the symptomatologies of ODD and OCD did not significantly contribute to SampEn alterations. Hence, reduced SampEn observed in the whole ADHD sample is primarily driven by ADHD symptomatology rather than the severity of comorbidities. So far, only two fMRI studies compared ADHD comorbid with ODD with comorbid-free ADHD, and findings indicated that the comorbid ADHD was associated with greater impairment in brain function than ADHD alone in the regions associated with EF, which aligned with the current observation: Noordemeer^41^ reported ADHD comorbid with ODD revealed reduced activity in the paracingulate and superior frontal gyrus during failed inhibition in the inhibition task. Wang et al.^42^ reported enhanced resting-state functional connectivity in ADHD comorbid with ODD in the regions including the superior/middle/medial frontal gyrus and middle cingulate gyrus using the bilateral anterior cerebellum as the seed.

Several caveats should be considered with respect to the current results. First, in our analysis of comorbid ADHD, ODD, and OCD, the model assumed that SampEn depended on a linear combination of ADHD, ODD, OCD, and other symptomatologies but not their interactions. To build a feasible model that incorporates interaction terms, it would be necessary to include individuals with ODD and/or OCD but without ADHD. We focused on the changes of complexity when ADHD coexisted with ODD, OCD, and other conditions. Including subjects without ADHD but with other psychiatric diagnoses would deviate from this objective, so we opted for a simpler model without interaction terms. Second, Sokunbi et al.^24^ reported a negative association between SampEn and the ADHD score in multiple regions, including the medial frontal gyrus. The current study did not find a significant association between complexity and ADHD, ODD, and OCD symptom severity. Third, we found the medicated ADHD pre-adolescents did not have significantly different SampEn than the non-medicated ADHD pre-adolescents.

In summary, our study demonstrated the importance of accounting for comorbid status when investigating neuropsychiatric disorders. There is substantial overlap in brain areas involved in cognitive and behavioral deficits observed across a range of psychiatric disorders, like ADHD and ODD, leading to some support to the notion of shared and overlapping mechanisms. When ADHD co-occurred with other psychiatric disorders, the reduction in SampEn extended beyond the regions affected in comorbid-free ADHD, indicating that comorbidities amplify neural complexity deficits. However, the symptomatology of ODD did not appear to impact the complexity value when co-occurring with ADHD. These findings highlight the need to consider the differential neural impacts of comorbid conditions and suggest the importance of further research to identify biomarkers that may inform targeted interventions.

## Data Availability

ABCD study

## Acknowledgments

Research reported in this publication was supported by the Office Of The Director, National Institutes Of Health of the National Institutes of Health under Award Numbers R01-AG066711 and S10OD032285. The content is solely the responsibility of the authors and does not necessarily represent the official views of the National Institutes of Health.

Data used in the preparation of this article were obtained from the Adolescent Brain Cognitive DevelopmentSM (ABCD) Study (https://abcdstudy.org), held in the NIMH Data Archive (NDA). This is a multisite, longitudinal study designed to recruit more than 10,000 children age 9-10 and follow them over 10 years into early adulthood. The ABCD Study® is supported by the National Institutes of Health and additional federal partners under award numbers U01DA041048, U01DA050989, U01DA051016, U01DA041022, U01DA051018, U01DA051037, U01DA050987, U01DA041174, U01DA041106, U01DA041117, U01DA041028, U01DA041134, U01DA050988, U01DA051039, U01DA041156, U01DA041025, U01DA041120, U01DA051038, U01DA041148, U01DA041093, U01DA041089, U24DA041123, U24DA041147. A full list of supporters is available at https://abcdstudy.org/federal-partners.html. A listing of participating sites and a complete listing of the study investigators can be found at https://abcdstudy.org/consortium_members/. ABCD consortium investigators designed and implemented the study and/or provided data but did not necessarily participate in the analysis or writing of this report. This manuscript reflects the views of the authors and may not reflect the opinions or views of the NIH or ABCD consortium investigators.

## Notes

### Competing Interest Statement

The authors have declared no competing interest.

### Funding Statement

NIH R01-AG066711 & S10OD032285

## References

(1) American Psychiatric Association.; American Psychiatric Association. DSM-5 Task Force. Diagnostic and statistical manual of mental disorders : DSM-5; American Psychiatric Association, 2013.

(2) Kessler, R. C.; Adler, L.; Barkley, R.; Biederman, J.; Conners, C. K.; Demler, O.; Faraone, S. V.; Greenhill, L. L.; Howes, M. J.; Secnik, K.;, et al. The prevalence and correlates of adult ADHD in the United States: results from the National Comorbidity Survey Replication. Am J Psychiatry 2006, 163 (4), 716–723. DOI: 10.1176/ajp.2006.163.4.716.

(3) QuickStats: Percentage* of Children and Adolescents Aged 5-17 Years Who Had Ever Received a Diagnosis of Attention-Deficit/Hyperactivity Disorder,. MMWR Morb Mortal Wkly Rep 2024, 73 (5), 116. DOI: 10.15585/mmwr.mm7305a6.

(4) Salehinejad, M. A.; Ghanavati, E.; Rashid, M. H. A.; Nitsche, M. A. Hot and cold executive functions in the brain: A prefrontal-cingular network. Brain Neurosci Adv 2021, 5, 23982128211007769. DOI: 10.1177/23982128211007769.

(5) Noordermeer, S. D.; Luman, M.; Oosterlaan, J. A Systematic Review and Meta-analysis of Neuroimaging in Oppositional Defiant Disorder (ODD) and Conduct Disorder (CD) Taking Attention-Deficit Hyperactivity Disorder (ADHD) Into Account. Neuropsychol Rev 2016, 26 (1), 44–72. DOI: 10.1007/s11065-015-9315-8.

(6) Rubia, K. "Cool" inferior frontostriatal dysfunction in attention-deficit/hyperactivity disorder versus "hot" ventromedial orbitofrontal-limbic dysfunction in conduct disorder: a review. Biol Psychiatry 2011, 69 (12), e69–87. DOI: 10.1016/j.biopsych.2010.09.023.

(7) Picó-Pérez, M.; Moreira, P. S.; de Melo Ferreira, V.; Radua, J.; Mataix-Cols, D.; Sousa, N.; Soriano-Mas, C.; Morgado, P. Modality-specific overlaps in brain structure and function in obsessive-compulsive disorder: Multimodal meta-analysis of case-control MRI studies. Neurosci Biobehav Rev 2020, 112, 83–94. DOI: 10.1016/j.neubiorev.2020.01.033.

(8) McDonough, I. M.; Nashiro, K. Network complexity as a measure of information processing across resting-state networks: evidence from the Human Connectome Project. Front Hum Neurosci 2014, 8, 409. DOI: 10.3389/fnhum.2014.00409.

(9) Smith, R. X.; Yan, L.; Wang, D. J. Multiple time scale complexity analysis of resting state FMRI. Brain Imaging Behav 2014, 8 (2), 284–291. DOI: 10.1007/s11682-013-9276-6.

(10) Wang, D. J. J.; Jann, K.; Fan, C.; Qiao, Y.; Zang, Y. F.; Lu, H.; Yang, Y. Neurophysiological Basis of Multi-Scale Entropy of Brain Complexity and Its Relationship With Functional Connectivity. Front Neurosci 2018, 12, 352. DOI: 10.3389/fnins.2018.00352.

(11) Wang, Z.; Li, Y.; Childress, A. R.; Detre, J. A. Brain entropy mapping using fMRI. PLoS One 2014, 9 (3), e89948. DOI: 10.1371/journal.pone.0089948.

(12) Fernández, A.; Gómez, C.; Hornero, R.; López-Ibor, J. J. Complexity and schizophrenia. Prog Neuropsychopharmacol Biol Psychiatry 2013, 45, 267–276. DOI: 10.1016/j.pnpbp.2012.03.015.

(13) Sun, J.; Wang, B.; Niu, Y.; Tan, Y.; Fan, C.; Zhang, N.; Xue, J.; Wei, J.; Xiang, J. Complexity Analysis of EEG, MEG, and fMRI in Mild Cognitive Impairment and Alzheimer’s Disease: A Review. Entropy (Basel) 2020, 22 (2). DOI: 10.3390/e22020239.

(14) Takahashi, T. Complexity of spontaneous brain activity in mental disorders. Prog Neuropsychopharmacol Biol Psychiatry 2013, 45, 258–266. DOI: 10.1016/j.pnpbp.2012.05.001.

(15) Richman, J. S.; Moorman, J. R. Physiological time-series analysis using approximate entropy and sample entropy. Am J Physiol Heart Circ Physiol 2000, 278 (6), H2039–2049. DOI: 10.1152/ajpheart.2000.278.6.H2039.

(16) Friston, K. J. Functional and effective connectivity: a review. Brain Connect 2011, 1 (1), 13–36. DOI: 10.1089/brain.2011.0008.

(17) Zang, Y. F.; He, Y.; Zhu, C. Z.; Cao, Q. J.; Sui, M. Q.; Liang, M.; Tian, L. X.; Jiang, T. Z.; Wang, Y. F. Altered baseline brain activity in children with ADHD revealed by resting-state functional MRI. Brain Dev 2007, 29 (2), 83–91. DOI: 10.1016/j.braindev.2006.07.002.

(18) Zang, Y.; Jiang, T.; Lu, Y.; He, Y.; Tian, L. Regional homogeneity approach to fMRI data analysis. Neuroimage 2004, 22 (1), 394–400. DOI: 10.1016/j.neuroimage.2003.12.030.

(19) Bullmore, E.; Sporns, O. Complex brain networks: graph theoretical analysis of structural and functional systems. Nat Rev Neurosci 2009, 10 (3), 186–198. DOI: 10.1038/nrn2575.

(20) Smith, S. M.; Fox, P. T.; Miller, K. L.; Glahn, D. C.; Fox, P. M.; Mackay, C. E.; Filippini, N.; Watkins, K. E.; Toro, R.; Laird, A. R.;, et al. Correspondence of the brain’s functional architecture during activation and rest. Proc Natl Acad Sci U S A 2009, 106 (31), 13040–13045. DOI: 10.1073/pnas.0905267106.

(21) Watts, D. J.; Strogatz, S. H. Collective dynamics of ’small-world’ networks. Nature 1998, 393 (6684), 440–442. DOI: 10.1038/30918.

(22) Meunier, D.; Lambiotte, R.; Bullmore, E. T. Modular and hierarchically modular organization of brain networks. Front Neurosci 2010, 4, 200. DOI: 10.3389/fnins.2010.00200.

(23) Guan, S.; Wan, D.; Zhao, R.; Canario, E.; Meng, C.; Biswal, B. B. The complexity of spontaneous brain activity changes in schizophrenia, bipolar disorder, and ADHD was examined using different variations of entropy. Hum Brain Mapp 2023, 44 (1), 94–118. DOI: 10.1002/hbm.26129.

(24) Sokunbi, M. O.; Fung, W.; Sawlani, V.; Choppin, S.; Linden, D. E.; Thome, J. Resting state fMRI entropy probes complexity of brain activity in adults with ADHD. Psychiatry Res 2013, 214 (3), 341–348. DOI: 10.1016/j.pscychresns.2013.10.001.

(25) Zhang, R.; Murray, S. B.; Duval, C. J.; Wang, D. J. J.; Jann, K. Functional connectivity and complexity analyses of resting-state fMRI in pre-adolescents demonstrating the behavioral symptoms of ADHD. Psychiatry Res 2024, 334, 115794. DOI: 10.1016/j.psychres.2024.115794.

(26) Jiang, X.; Li, X.; Xing, H.; Huang, X.; Xu, X.; Li, J. Brain Entropy Study on Obsessive-Compulsive Disorder Using Resting-State fMRI. Front Psychiatry 2021, 12, 764328. DOI: 10.3389/fpsyt.2021.764328.

(27) Aydin, S.; Arica, N.; Ergul, E.; Tan, O. Classification of obsessive compulsive disorder by EEG complexity and hemispheric dependency measurements. Int J Neural Syst 2015, 25 (3), 1550010. DOI: 10.1142/S0129065715500100.

(28) Yazdi-Ravandi, S.; Mohammadi Arezooji, D.; Matinnia, N.; Shamsaei, F.; Ahmadpanah, M.; Ghaleiha, A.; Khosrowabadi, R. Complexity of information processing in obsessive-compulsive disorder based on fractal analysis of EEG signal. EXCLI J 2021, 20, 462–654. DOI: 10.17179/excli2020-2783.

(29) Casey, B. J.; Cannonier, T.; Conley, M. I.; Cohen, A. O.; Barch, D. M.; Heitzeg, M. M.; Soules, M. E.; Teslovich, T.; Dellarco, D. V.; Garavan, H.;, et al. The Adolescent Brain Cognitive Development (ABCD) study: Imaging acquisition across 21 sites. Dev Cogn Neurosci 2018, 32, 43–54. DOI: 10.1016/j.dcn.2018.03.001.

(30) Roediger, D. J.; Butts, J.; Falke, C.; Fiecas, M. B.; Klimes-Dougan, B.; Mueller, B. A.; Cullen, K. R. Optimizing the measurement of sample entropy in resting-state fMRI data. Front Neurol 2024, 15, 1331365. DOI: 10.3389/fneur.2024.1331365.

(31) Desikan, R. S.; Ségonne, F.; Fischl, B.; Quinn, B. T.; Dickerson, B. C.; Blacker, D.; Buckner, R. L.; Dale, A. M.; Maguire, R. P.; Hyman, B. T.;, et al. An automated labeling system for subdividing the human cerebral cortex on MRI scans into gyral based regions of interest. Neuroimage 2006, 31 (3), 968–980. DOI: 10.1016/j.neuroimage.2006.01.021.

(32) Benjamini, Y.; Hochberg, Y. Controlling the false discovery rate: a practical and powerful approach to multiple testing. Journal of the Royal Statistical Society. Series B (Methodological) 1995, 57 (1), 12.

(33) Achenback, T. M.; Rescorla, L. A. Manual for the ASEBA School-Age Forms & Profiles. ASEBA: Burlington, VT, 2001.

(34) Achenbach, T. M.; Rescorla, L. A. Multicultural Supplement to the Manual for the ASEBA School-Age Forms & Profiles. ASEBA: Burlington, VT, 2007.

(35) Liu, N.; Liu, Q.; Yang, Z.; Xu, J.; Fu, G.; Zhou, Y.; Li, H.; Wang, Y.; Liu, L.; Qian, Q. Different functional alteration in attention-deficit/hyperactivity disorder across developmental age groups: A meta-analysis and an independent validation of resting-state functional connectivity studies. CNS Neurosci Ther 2023, 29 (1), 60–69. DOI: 10.1111/cns.14032.

(36) Gao, Y.; Shuai, D.; Bu, X.; Hu, X.; Tang, S.; Zhang, L.; Li, H.; Lu, L.; Gong, Q.; Huang, X. Impairments of large-scale functional networks in attention-deficit/hyperactivity disorder: a meta-analysis of resting-state functional connectivity. Psychol Med 2019, 49 (15), 2475–2485. DOI: 10.1017/S003329171900237X.

(37) Sutcubasi, B.; Metin, B.; Kurban, M. K.; Metin, Z. E.; Beser, B.; Sonuga-Barke, E. Resting-state network dysconnectivity in ADHD: A system-neuroscience-based meta-analysis. World J Biol Psychiatry 2020, 21 (9), 662–672. DOI: 10.1080/15622975.2020.1775889.

(38) Blair, R. J. R.; Veroude, K.; Buitelaar, J. K. Neuro-cognitive system dysfunction and symptom sets: A review of fMRI studies in youth with conduct problems. Neurosci Biobehav Rev 2018, 91, 69–90. DOI: 10.1016/j.neubiorev.2016.10.022.

(39) Bruin, W.; Denys, D.; van Wingen, G. Diagnostic neuroimaging markers of obsessive-compulsive disorder: Initial evidence from structural and functional MRI studies. Prog Neuropsychopharmacol Biol Psychiatry 2019, 91, 49–59. DOI: 10.1016/j.pnpbp.2018.08.005.

(40) Brem, S.; Hauser, T. U.; Iannaccone, R.; Brandeis, D.; Drechsler, R.; Walitza, S. Neuroimaging of cognitive brain function in paediatric obsessive compulsive disorder: a review of literature and preliminary meta-analysis. J Neural Transm (Vienna) 2012, 119 (11), 1425–1448. DOI: 10.1007/s00702-012-0813-z.

(41) Noordermeer, S. The ODD part of ADHD. Vrije Universiteit Amsterdam, 2019.

(42) Wang, X.; Guo, Y.; Xu, J.; Xiao, Y.; Fu, Y. Decreased gray matter volume in the anterior cerebellar of attention deficit/hyperactivity disorder comorbid oppositional defiant disorder children with associated cerebellar-cerebral hyperconnectivity: insights from a combined structural MRI and resting-state fMRI study. Int J Dev Neurosci 2024. DOI: 10.1002/jdn.10349.

(43) Petersen, A. C.; Crockett, L.; Richards, M.; Boxer, A. A self-report measure of pubertal status: Reliability, validity, and initial norms. J Youth Adolesc 1988, 17 (2), 117–133. DOI: 10.1007/BF01537962.

