## Supplementary for "Elucidating distinct and common fMRI-complexity patterns in pre-adolescent children with Attention-Deficit/Hyperactivity Disorder, Oppositional Defiant Disorder, and Obsessive-Compulsive Disorder diagnoses"

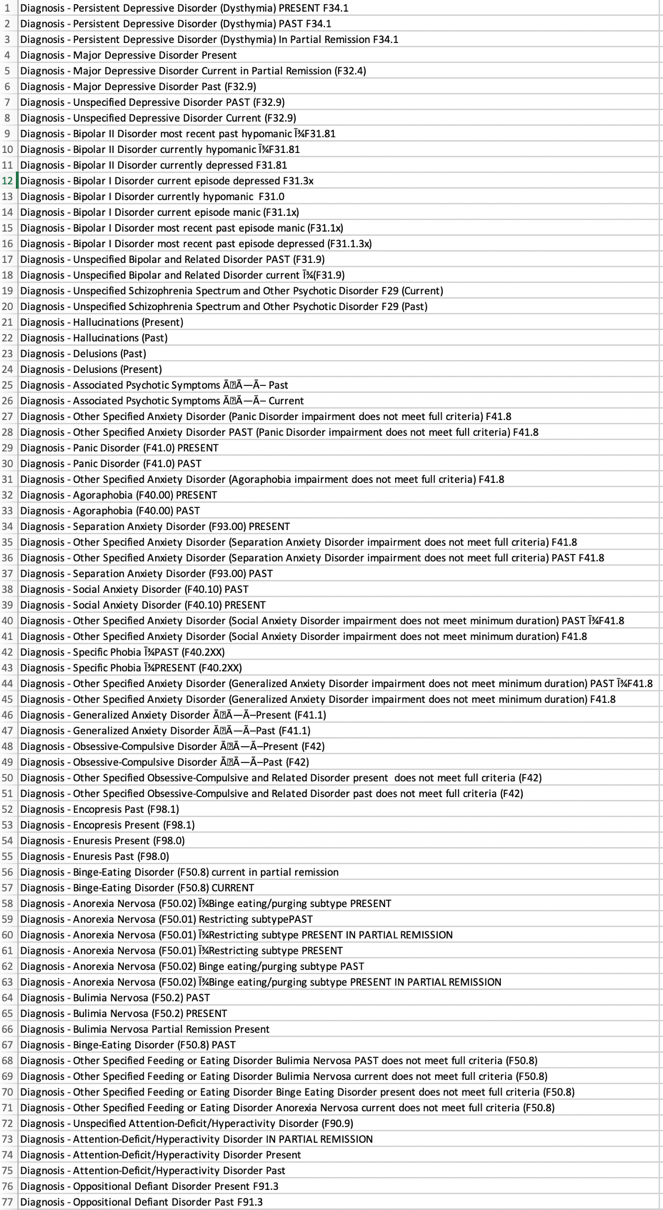

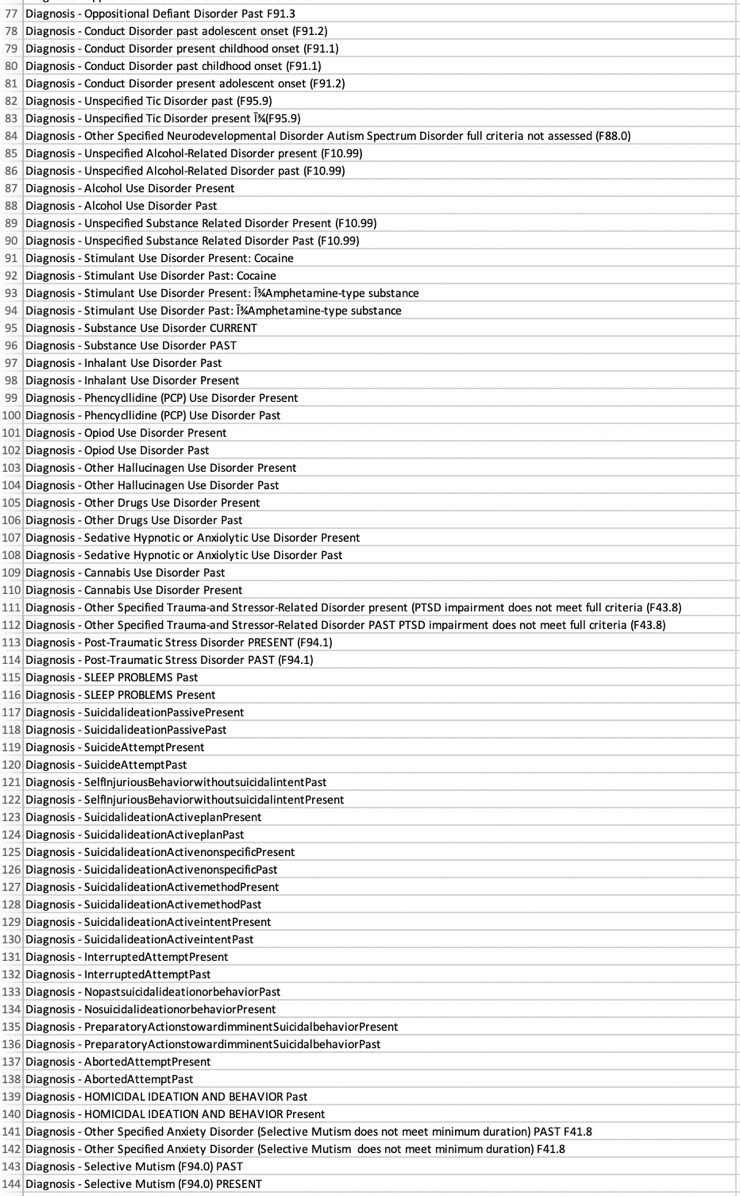

Figure S1. 144 psychiatric diagnoses evaluated by the ABCD study.

More details about the diagnosis of ADHD in ABCD release 3.0

In release 3.0, ADHD diagnoses were made according to DSM-5 criteria,^1^ according to parent reports by the Kiddie-Schedule for Affective Disorders and Schizophrenia (KSADS)-parent report. 1087 baseline participants were diagnosed with ADHD (i.e., Attention-Deficit/Hyperactivity Disorder Present) with or without various comorbidities, comprising 9.2% of the entire ABCD baseline sample. This aligns with previous studies, which have reported that ADHD affects approximately 11.3% of children and adolescents in the United States.^2^ However, due to an anomaly within the ABCD Study infrastructure, what was originally confirmed to represent ADHD diagnoses was subsequently found to be invalid, on the grounds that parents were asked whether “symptoms interfere with social, academic or occupational functioning”. Full DSM-5 criteria mandate that the impairment caused by symptoms extends to at least two domains. As such, full threshold DSM-5 diagnoses could not be confirmed, and our sample here comprised subjects who met all the behavioral criteria for ADHD diagnoses, without knowing whether the impairment extended to multiple domains. Therefore, our sample related to children with all behavioral features of ADHD.

Imaging Acquisition and Preprocessing

All T1-weighted scans were acquired with voxel resolution = 1mm^3^, 256 × 256 matrix, flip angle = 8°, and 2x parallel imaging. Other scan parameters slightly varied by scanner platform, i.e., Siemens Prisma, Philips, or GE 3T scanner. Each participant had 3-4 eyes-open (passive crosshair viewing) rsfMRI scans, each of which was approximately 5-minutes in duration. All rsfMRI scans were collected using a gradient-echo EPI sequence of 383 volumes in total (voxel resolution = $2.4\times2.4\times2.4$ mm^3^, 60 slices, 90 × 90 matrix, FOV = 216 × 216, TR = 800 ms, TE = 30 ms, flip angle = 52°, 6-factor multiband acceleration). For the current study, we used rsfMRI run 1 and run 2. For each subject T1 structural and resting-state fMRI data were preprocessed using CONN toolbox (Conn: fMRI functional connectivity toolbox) including motion realignment, regression of motion and physiological noise-related signal variations, coregistration and normalization of structural and functional scans to standard MNI space, and spatial smoothing with a 6mm FWHM Gaussian Kernel.

Clinical Results

Previous studies indicated the comorbidity rate of ODD within ADHD children and adolescents is around 26% and the rate of co-occurrence of OCD ranges from 3% to 7.5%.^3, 4^ In the current ADHD sample, the probability of being diagnosed as ODD was 30%, consistent with published rates, whereas the rate of being diagnosed as OCD was 22.5%, notably higher than expected. Independent-sample *t*-tests indicated that the ADHD score of the individuals with ADHD plus various comorbidities was significantly higher than those with comorbid-free ADHD (Mean difference of the ADHD score = 2.475, *t*(892) = 6.514, *p* < 0.05); ODD score of the individuals with ODD plus various comorbidities was significantly higher than those with comorbid-free ODD (Mean difference of the ODD score = 2.916, *t*(304) = 8.060, *p* < 0.05); OCD score of the individuals with OCD plus various comorbidities was significantly higher than those with comorbid-free OCD (Mean difference of the OCD score = 3.149, *t*(247) = 7.317, *p* < 0.05); It suggested that symptom severity is greater when it co-occurs with other symptoms compared to when it presents alone, which was consistent with observations in literature.^5-7^

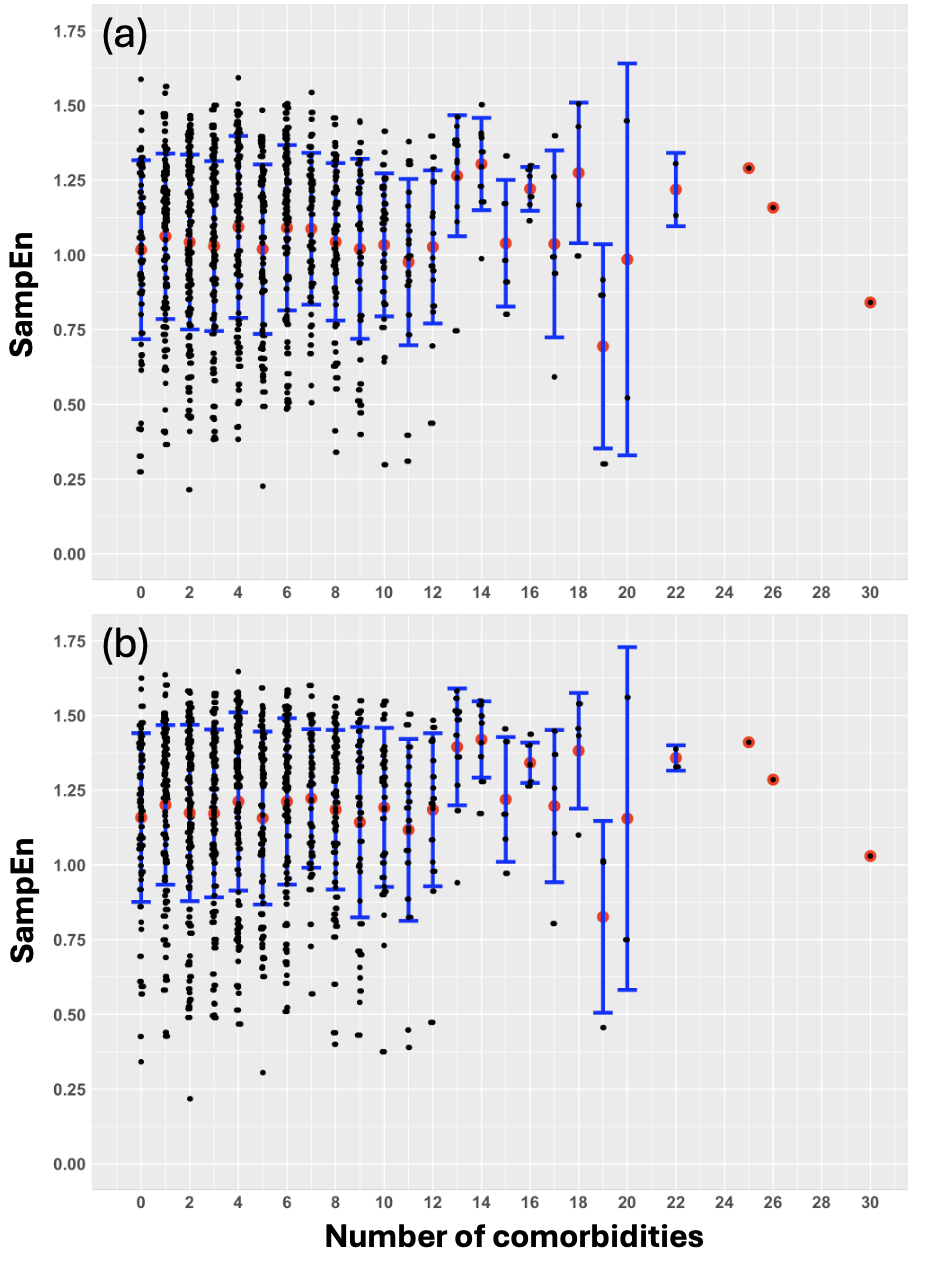

Figure S2. (a) Mean SampEn of the cool EF network for each ADHD subject, plotted against the number of comorbidities. The red dot and blue line segment display the mean and standard deviation of each condition. (b) Mean SampEn of the hot EF network for each ADHD subject, plotted against the number of comorbidities. The red dot and blue line segment display the mean and standard deviation of each condition.

Table S1. A list of ADHD prescriptions.

Prescription

Adderall

Azstarys

Serdexmethylphenidate

Dexmethylphendiate

Concerta

Methylphenidate

Focalin

Intuniv

Guanfacine

Gelbree

Viloxazine

Ritalin

Strattera

Atomoxetine

Vyvanse

Adzenys

Evekeo

Daytrana

Quillivant

Amphetamine

Clonidine

Metadate

References

(1) American Psychiatric Association.; American Psychiatric Association. DSM-5 Task Force. *Diagnostic and statistical manual of mental disorders : DSM-5*; American Psychiatric Association, 2013.

(2) QuickStats: Percentage* of Children and Adolescents Aged 5-17 Years Who Had Ever Received a Diagnosis of Attention-Deficit/Hyperactivity Disorder,. *MMWR Morb Mortal Wkly Rep* **2024**, *73* (5), 116. DOI: 10.15585/mmwr.mm7305a6.

(3) Abramovitch, A.; Dar, R.; Mittelman, A.; Wilhelm, S. Comorbidity Between Attention Deficit/Hyperactivity Disorder and Obsessive-Compulsive Disorder Across the Lifespan: A Systematic and Critical Review. *Harv Rev Psychiatry* **2015**, *23* (4), 245-262. DOI: 10.1097/HRP.0000000000000050.

(4) Mohammadi, M. R.; Zarafshan, H.; Khaleghi, A.; Ahmadi, N.; Hooshyari, Z.; Mostafavi, S. A.; Ahmadi, A.; Alavi, S. S.; Shakiba, A.; Salmanian, M. Prevalence of ADHD and Its Comorbidities in a Population-Based Sample. *J Atten Disord* **2021**, *25* (8), 1058-1067. DOI: 10.1177/1087054719886372.

(5) Azeredo, A.; Moreira, D.; Barbosa, F. ADHD, CD, and ODD: Systematic review of genetic and environmental risk factors. *Res Dev Disabil* **2018**, *82*, 10-19. DOI: 10.1016/j.ridd.2017.12.010.

(6) Steiner, H.; Remsing, L.; Issues, W. G. o. Q. Practice parameter for the assessment and treatment of children and adolescents with oppositional defiant disorder. *J Am Acad Child Adolesc Psychiatry* **2007**, *46* (1), 126-141. DOI: 10.1097/01.chi.0000246060.62706.af.

(7) Walitza, S.; Zellmann, H.; Irblich, B.; Lange, K. W.; Tucha, O.; Hemminger, U.; Wucherer, K.; Rost, V.; Reinecker, H.; Wewetzer, C.; et al. Children and adolescents with obsessive-compulsive disorder and comorbid attention-deficit/hyperactivity disorder: preliminary results of a prospective follow-up study. *J Neural Transm (Vienna)* **2008**, *115* (2), 187-190. DOI: 10.1007/s00702-007-0841-2.
